# Predicting vasospasm risk using first presentation aneurysmal subarachnoid haemorrhage volume: a semi-automated CT image segmentation analysis in ITK-SNAP

**DOI:** 10.1101/2023.01.22.23284860

**Authors:** James S Street, Anand S Pandit, Ahmed K Toma

**Affiliations:** Medical School, Faculty of Medical Sciences, University College London, London, United Kingdom; Victor Horsley Department of Neurosurgery, The National Hospital for Neurology and Neurosurgery, Queen Square, London, United Kingdom; High-Dimensional Neurology, Institute of Neurology, University College London, London, United Kingdom

**Keywords:** Subarachnoid haemorrhage, Vasospasm, Machine learning, Image segmentation

## Abstract

**Purpose:** Cerebral vasospasm following aneurysmal subarachnoid haemorrhage (aSAH) is a significant complication associated with poor neurological outcomes. We present a novel, semi-automated pipeline in ITK-SNAP to segment subarachnoid blood volume from initial CT head (CTH) scans and use this to predict future radiological vasospasm.

**Methods:** 42 patients were admitted between February 2020 and December 2021 to our tertiary neurosciences centre, and whose initial referral CTH scan was used for this retrospective cohort study. Blood load was segmented using a semi-automated random forest classifier and active contour evolution implemented in the open-source medical imaging analysis software ITK-SNAP. Clinical data were extracted from electronic healthcare records in order to fit models aimed at predicting radiological vasospasm risk.

**Results:** Semi-automated segmentations demonstrated excellent agreement with manual, expert-derived volumes (mean Dice coefficient=0.92). Total normalised blood volume, extracted from CTH images at first presentation, was significantly associated with greater odds of later radiological vasospasm, increasing by approximately 7% for each additional cm^3^ of blood (OR=1.069, 95% CI: 1.021-1.120; p<.005). Greater blood volume was also significantly associated with vasospasm of a higher Lindegaard ratio, of longer duration, and a greater number of discrete episodes. Total blood volume predicted radiological vasospasm with a greater accuracy as compared to the modified Fisher scale (AUC= 0.86 vs 0.70), and was of independent predictive value.

**Conclusion:** Semi-automated methods provide a plausible pipeline for the segmentation of blood from CT head images in aSAH, and total blood volume is a robust, extendable predictor of radiological vasospasm, outperforming the modified Fisher scale. Greater subarachnoid blood volume significantly increases the odds of subsequent vasospasm, its time course and its severity.

## Introduction

Outcomes following aneurysmal subarachnoid haemorrhage (aSAH) remain poor, with an estimated mortality of approximately 30%[1]. An important contributor to both morbidity and mortality following aSAH is cerebral vasospasm, the spasmodic narrowing of intracranial arteries which can lead to delayed cerebral ischaemia (DCI). Symptomatic vasospasm occurs in 20% of patients[2], and typically occurs at a delay of 3 to 12 days after the haemorrhagic event[3]. Whereas angiographic or radiologically-detected vasospasm can be detected in as many as 50-70% of aSAH patients, not all are associated with neurological deficits. As such, the prediction and forward recognition of clinically significant vasospasm represents a substantial challenge in the management of these patients.

One predictor of future vasospasm is the total volume of blood seen on the CT head scan at presentation[4–8]. Furthermore, intraventricular haemorrhage (IVH) has been shown to independently predict cerebral vasospasm[6,8–13], and total blood volume is associated with worse functional outcomes[14]. Drawing on these key findings, aSAH severity is frequently graded in clinical practice using the modified Fisher scale (mFS)[8] -- a subjective assessment of bleed extent on CT head scans. However, the modified Fisher scale only crudely notes blood distribution and blood load, and accordingly, its qualitative nature limits its predictive power. Troublingly, the modified Fisher scale has recently been demonstrated to lack inter-rater reliability[15], and thus may not provide an objective metric of blood burden and distribution following aSAH.

Further work has introduced similar qualitative or semi-quantitative severity scores for the purposes of predicting vasospasm from cisternal blood volume[5], intraventricular blood volume[16], and intraparenchymal blood volume[17]. Yet, these scales are also observer-dependent and can exhibit poor inter-rater agreement[18,19]. Nevertheless, in patients of poor clinical grade whose clinical neurology is difficult to assess, these metrics represent some of the few available clinical tools which can inform the likelihood of vasospasm and guide risk-stratification and management. The precise relationship between quantified blood volume and radiological vasospasm likelihood remains poorly defined.

Although the presence and degree of blood in cerebral compartments is associated with clinical, symptomatic vasospasm[8,12,19], little work has looked at the prediction of radiological vasospasm from routinely acquired neuroimaging data. Although the precise relationship between radiological vasospasm and DCI is contested[20], there is strong recent evidence of its association with clinical and functional outcomes [21,22]. As the onset of DCI is difficult to diagnose and frequently missed [23], such radiological outcomes remain important to guide decisions regarding clinical intervention, including angioplasty [24].

Quantitative image segmentation approaches, such as those used to segment other organ systems[25], can detail both the volume and morphology of blood load and would overcome the aforementioned difficulties, potentially offering a more accurate method for vasospasm prediction. However, manual image segmentation of haemorrhage on CT head scans can be technically challenging, time-consuming and impractical for clinical use. Novel developments in medical image segmentation mean that precise and robust estimates of blood volume and distribution are easier to calculate. These include semi-automated tools such as active contour evolution and clustering based algorithms[26] which are more efficient and have precedent in delineating vascular structures in other areas[27].

Here, we present a working pipeline, implemented in ITK-SNAP, for the efficient, semi-automated segmentation of blood on a non-contrast, routine plain CT head scan, obtained at first presentation of patients with aSAH. We demonstrate that semi-automated segmentations align well with clinical expert impressions of blood distribution and require minimal correction. Our model is compared against the current standard of the modified Fisher scale in correlating against occurrence of any radiological vasospasm as the primary outcome. Secondary outcomes included time to, duration and number of discrete vasospasm episodes and general reported outcome measures of length of intensive therapy unit (ITU) stay, hospital stay and mortality.

## Methods

### Protocol

The study was performed in accordance with the STROBE checklist[28] and the European Society of Radiology statement on imaging biomarkers[29] where relevant.

### Ethics

This retrospective cohort study was approved by the institutional review board (53-202122-CA) in the context of a wider service evaluation regarding radiological assessment of vasospasm in patients with aSAH at our tertiary neurosciences centre.

### Participants

A list of candidate patients were identified from the institutional electronic healthcare record (EHR) Patients were included in this study if they (i) were treated for an aneurysmal subarachnoid haemorrhage at the academic neurosciences centre between February 2020 - December 2021; (ii) the initial CT head (CTH) scan performed at first presentation was available on the clinical imaging repository; and (iii) the patient had no prior medical history of aSAH or intracranial haemorrhage, no previous intracranial coil embolisation, or any other intracranial implant in situ that would degrade CT image quality.

### Image processing and segmentation pipeline

The steps regarding image pre- and post-processing and haemorrhage segmentation, alongside the software packages used have been outlined in Figure 1. Briefly, the first CT head (CTH) scan performed following the SAH ictus was obtained from the Picture Archiving and Communication System (PACS). DICOM CT files were anonymised and converted to NIfTI format using the command line tool *dcm2niix[30]*. *dcm2niix* includes an inbuilt Gantry tilt correction routine that was used to ensure NIfTI files were correctly oriented. NIfTI files were loaded into the FMRIB software library (FSL)[31] and a binary mask image was generated to delineate the brain using FSL’s Brain Extraction Tool (BET). To perform BET, images were initially smoothed using a Gaussian kernel of size 1mm^3^ and thresholded between 0 to 120 Hounsfield Units (HU)[32] and parameterised for optimal extraction of brain tissue from CT images[32]. The extracted brain image was binarised to generate a brain mask that could be used in subsequent analysis, and was manually inspected for adequacy before being used.

**Fig 1.**
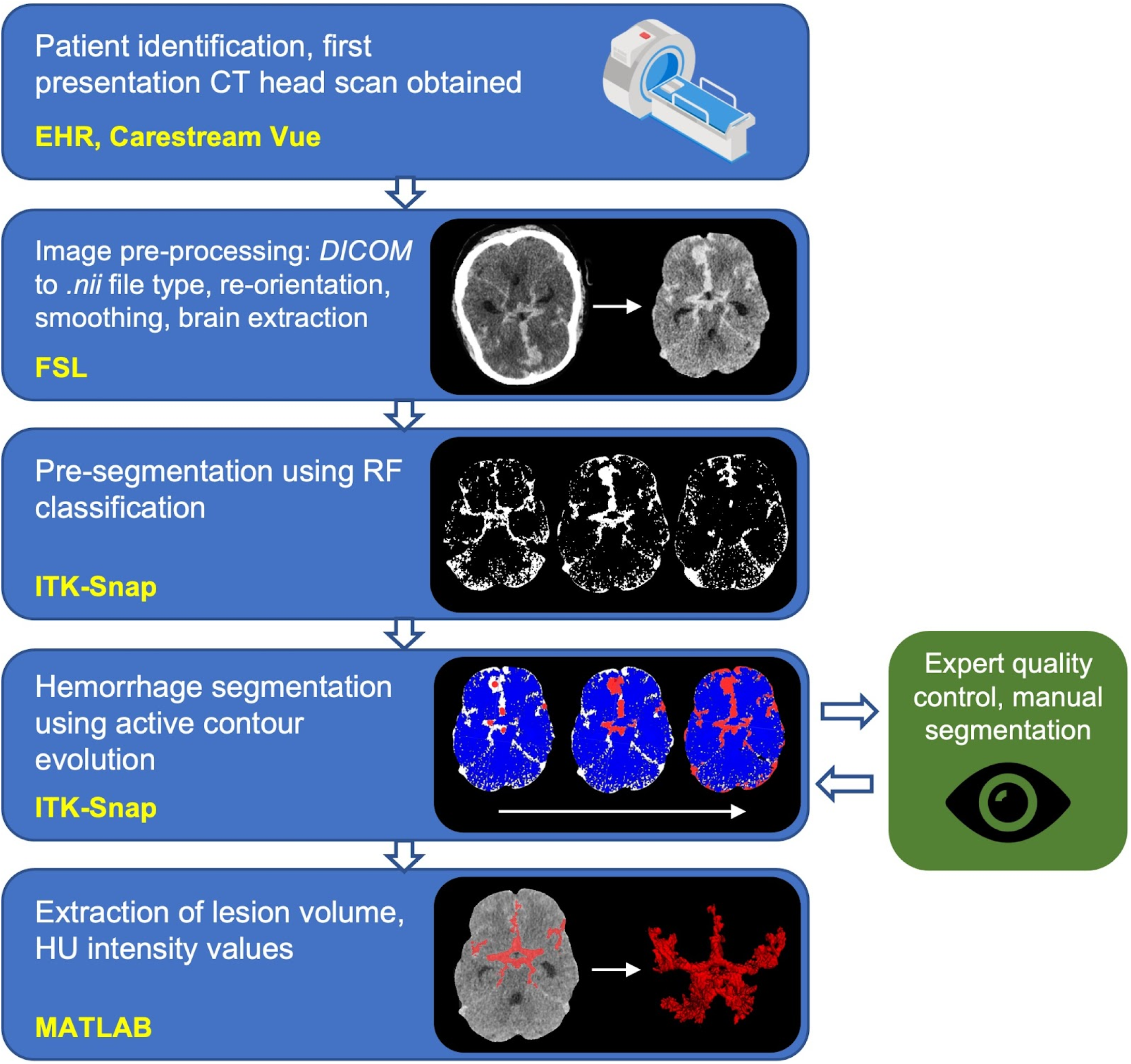
Image processing pipeline describing data collection, pre-processing, segmentation, quality control and information extraction. (yellow font = software library; DICOM = Digital Imaging and Communications in Medicine; nii = nifti file type; FSL = FMRIB Software Library; ITK-Snap = Insight Segmentation and Registration Toolkit; RF = random forest; HU = Hounsfield Units. ‘CT-scan’ designed using resources from Flatiron.com)

All segmentations were performed in ITK-SNAP, an open-source and multi-platform 3D medical image analysis software, optimised for user-guided segmentation[33]. All images were resampled using linear interpolation such that voxels were cubic in size to ensure consistency between patients and diverse scanner types. Briefly, to segment the image, a random forest classifier (tree depth = 30, number of trees = 50, classifier bias = 0.5) was initially trained on each image using manually labelled examples to classify voxels into one of four tissue subtypes: CSF, bone, parenchyma, or blood (Figure 1). HU values at each labelled voxel and the 2-voxel wide neighbourhood around each labelled voxel were used to train the classifier. This was used to generate a speed image, which encoded at each pixel the desired rate of growth or retraction of the contour. Spherical seeds were manually placed on the image to initialise the contour, which was then allowed to actively evolve over approximately 500 iterations. Segmentations took, on average, 12 minutes to perform for each brain. Example series of axial slices through four brains are shown in Figure 2, which display the segmentation output for all blood detected by this process.

**Fig 2:**
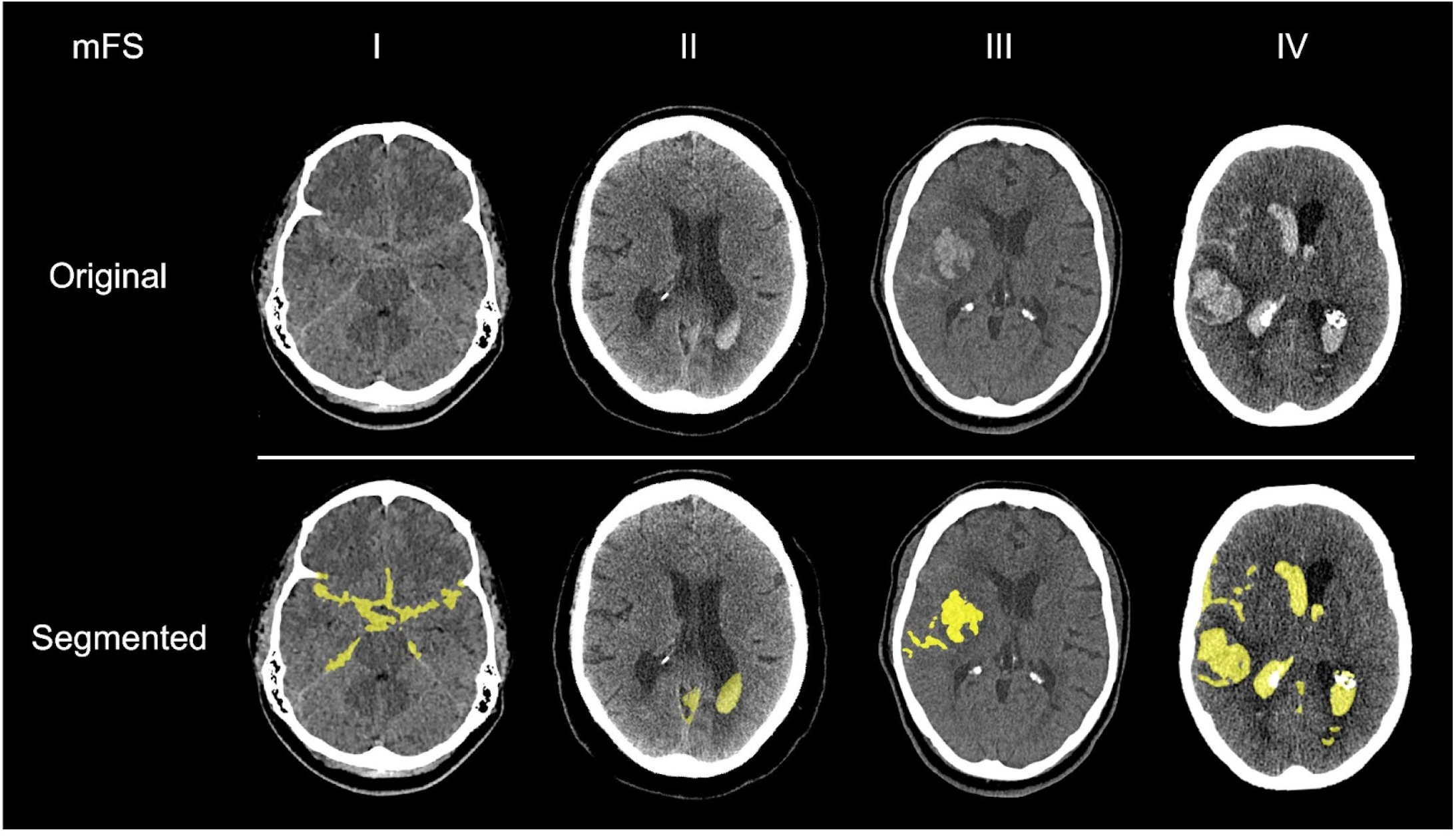
Examples of semi-automated subarachnoid haemorrhage segmentations for each modified Fisher grade. (yellow = segmentation overlay, mFS = modified Fisher Scale)

### Quality assessment

All segmentations were reviewed and manually corrected by an academic neurosurgical resident with a decade of post-doctoral neuroimaging experience (A.S.P.) and consultant neurovascular surgeon (A.K.T.) to provide finalised segmentations to act as the ground truth for subsequent analyses. Dice overlap coefficients were calculated between the original and expert-corrected segmentations to assess the quality of the semi-automated segmentation pipeline.

### Clinical data extraction

Relevant clinical data were extracted from the EHR and from radiology reports available on PACS. This data included, as the primary outcome, the presence of radiologically detectable vasospasm via: transcranial doppler (TCD), CT angiogram (CTA), or digital subtraction angiography (DSA). The majority of patients received repeat TCD imaging during recovery, with documented Lindegaard ratios (n=30), with CTA and DSA performed based on assessment of clinical need. Secondary outcomes included time to vasospasm, vasospasm duration (defined as the number of days with any positive radiological vasospasm), number of discrete vasospasm episodes (defined as the number of episodes of detected vasospasm that were separated by at least one day of exclusively negative tests), vasospasm severity, length of ITU admission, length of hospital admission, and mortality. Vasospasm severity was quantified in two ways. Firstly, the consultant neuroradiologist’s subjective impression of vessel calibre (using CTA or DSA) was extracted and categorised as: ‘none’, ‘mild’, ‘moderate’, or ‘severe’. Secondly, the greatest Lindegaard ratio was extracted from radiology records for all patients who received TCD imaging and where this was documented. Given that many of the patients were intubated and sedated during their initial hospital admission and the data was retrospectively collected, clinical vasospasm or delayed clinical ischaemia (DCI) could not be reliably ascertained. The modified Fisher grade was recorded by author A.S.P following manual review of the images.

### Data and statistical analysis

Generalised linear models were fitted in MATLAB (R2019b, Mathworks Inc., Natick, MA) to assess for significant associations between predictors and response variables. Where response variables were binary values, logistic regression was used, otherwise a linear regression model was used.

For logistic models, (McFadden’s) pseudo-R^2^, was calculated to estimate the model’s predictive power[34], with values between 0.2 - 0.4 representing an “excellent” model fit[35]. Model goodness-of-fit was calculated in reference to a null model fit with constant (intercept) terms alone. Additional comparisons between nested models were performed using the likelihood-ratio test, alongside evaluation of the Akaike information criterion (AIC) and Bayesian information criterion (BIC). Hosmer-Lemeshow and Stukel tests were used to assess for model fitting and misspecification (Supplementary Methods).

All values are reported as mean ± standard deviation, unless otherwise specified. Data were tested for normality using the one-sample Kolmogorov-Smirnov test. All significance tests, unless stated otherwise, were two-tailed with a significance threshold of 5%. Total blood volume was normalised and scaled *(total blood volume x mean participant brain volume / participant brain volume)* using the brain volume estimated from the BET images derived above. Blood volume reported is normalised to participant brain volume unless stated otherwise. The sample size was determined pragmatically, namely the maximum number of images obtained within the specified study period. Scatter plots, unless stated otherwise, are colour-coded according to the radiological vasospasm status of the patient (maroon: vasospasm detected by any modality during admission; blue: no vasospasm detected by any modality during admission).

## Results

### Participant demographics

Data from 42 patients were used for this study (see Table 1 for a summary of demographics). 71.4% (n=30) of the included patients developed radiological vasospasm, detected via TCD, CTA, or DSA during admission, comparable to previously reported rates of radiological vasospasm[3]. There was no evidence of a difference in vasospasm risk in patients following endovascular coil embolisation versus aneurysm clipping (χ^2^ = 1.45; p = 0.23), nor was vasospasm risk associated with patient gender (χ^2^ = 26; p = 0.61) or age (t = -1.00, p = 0.32). Patients with radiological vasospasm experienced similar ITU stays (360±202 hours vs 288±381 hours; p = 0.46), but remained in hospital for significantly longer as compared to those who did not (59.8±46.6 days vs 23.7±24.2 days; 95% CI [4.6, 67.6], t = 2.33, p = 0.026). Length of ITU stay was neither significantly correlated with the duration (r = 0.25; p = 0.14) or number of distinct vasospasm episodes (r = 0.09; p = 0.56).

**Table 1.** Descriptive information regarding each patient’s demographics, aneurysm, treatment and hospital outcomes. (*patient passed away before coil embolisation organised. AComm = anterior communicating artery, PComm = posterior communicating artery, MCA = middle cerebral artery, ICA = internal carotid artery, PICA = posterior inferior cerebellar artery, SCA = superior cerebellar artery)

|  |  |  |
| --- | --- | --- |
| **Number of patients** | 42 |  |
| **Mean age (SD)** | 57.8 (11.6) years |  |
| **Female sex (%)** | 27 (64.3) |  |
| **Aneurysmal location (%)** | AComm<br>PComm<br>MCA<br>ICA<br>PICA<br>SCA | 17 (40.5)<br>5 (11.9)<br>7 (16.7)<br>5 (11.9)<br>6 (14.3)<br>2 (4.8) |
| **Radiological vasospasm frequency determined by (%)** | TCD<br>CTA<br>DSA<br>Any modality | 23 (54.8)<br>30 (71.4)<br>16 (38.1)<br>30 (71.4) |
| **Treatment frequency:**<br>**CSF diversion (%)** | External ventricular drain | 30 (71.4) |
|  | Lumbar drain | 11 (26.2) |
|  | No CSF diversion | 11 (26.2) |
| **Treatment frequency:**<br>**Neurovascular (%)** | Coil embolisation | 32 (76.2) |
|  | Aneurysmal clipping | 9 (21.4) |
|  | *Other | 1 (2.4) |
| **Mean length of ITU stay (SD)** | 343 (251) hours |  |
| **Mean hospital stay (SD)** | 50.3 (44.6) days |  |
| **Mortality (%)** | 4 (9.5) |  |

### Subarachnoid blood volume can be accurately and reliably segmented and estimated using a semi-automated pipeline

In general, all segmentations agreed with the expert-corrected segmentations (Dice coefficient: median = 0.994; mean = 0.920) with a mean volumetric error of (mean = 2.49±5.82 cm^3^).

### Segmented subarachnoid blood volume is associated with radiological vasospasm risk

Patients who developed radiological vasospasm had a significantly greater non-normalised blood load on the CT head scan at initial presentation (vasospasm mean blood volume = 60.3±30.5 cm^3^; non-vasospasm mean blood volume = 24.2±21.4 cm^3^, t = 3.74, p < .001; Figure 3).

**Fig 3:**
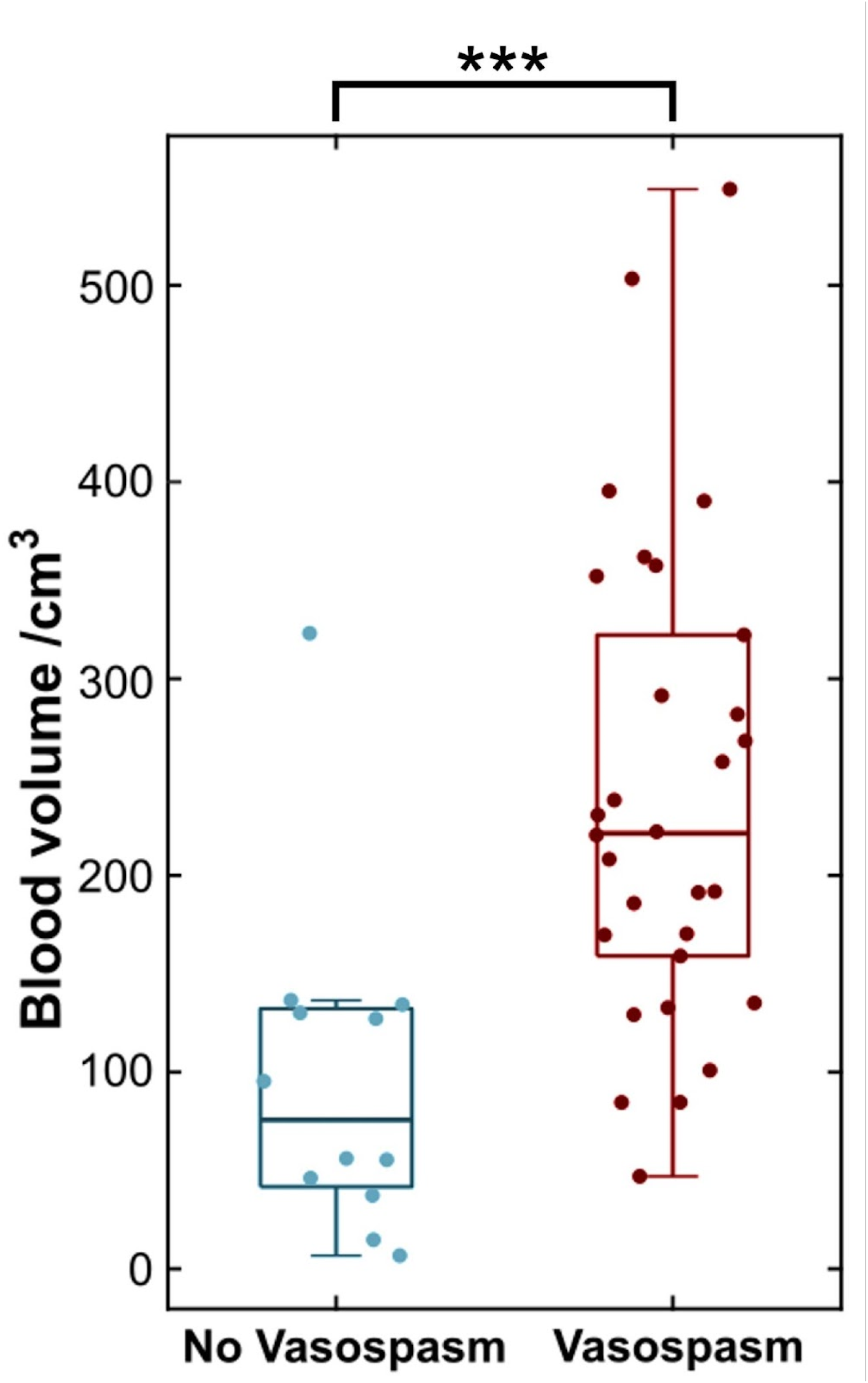
Greater segmented blood load is associated with greater radiological vasospasm risk. Boxplot of blood volume in patients who developed radiological vasospasm (maroon) and those who did not (blue). (*** = p< .001)

Similarly, logistic regression demonstrated a significant association between normalised blood volume and vasospasm risk (OR = 1.069 [95% CI: 1.021 - 1.120]; p = .0049; df = 40). This indicates that the odds of radiological vasospasm occurring during admission increase by approximately 7% for each cm^3^ of blood present in the subarachnoid spaces. This model explained a ‘good’ proportion of variance (pseudo-R^2^ = 0.30) and significantly more variance than a constant model (F = 15.2; p < .0001). There was no evidence that the model was misspecified (Stukel test: p_za_ = 0.61, p_zb_ = 0.51; Hosmer-Lemeshow test: χ^2^ = 3.49, df = 8, p = 0.90). A retriever operator characteristic (ROC) curve was constructed for this model (Figure 4). This demonstrated that the fitted model reliably separated the binary classes using normalised blood volume alone, and the ROC curve accordingly shows a high area-under-the-curve (AUC = 0.86). Furthermore, the significance of the relationship between blood volume and vasospasm was preserved after adding potential confounding variables into the model (see Table 2). Leave-one-out cross validation over this full model demonstrated a classification accuracy of 71.4% for subsequent vasospasm, with the full model maintaining a high predictive power (AUC = 0.89) and good proportion of explained variance (pseudo-R^2^ = 0.41).

**Fig 4:**
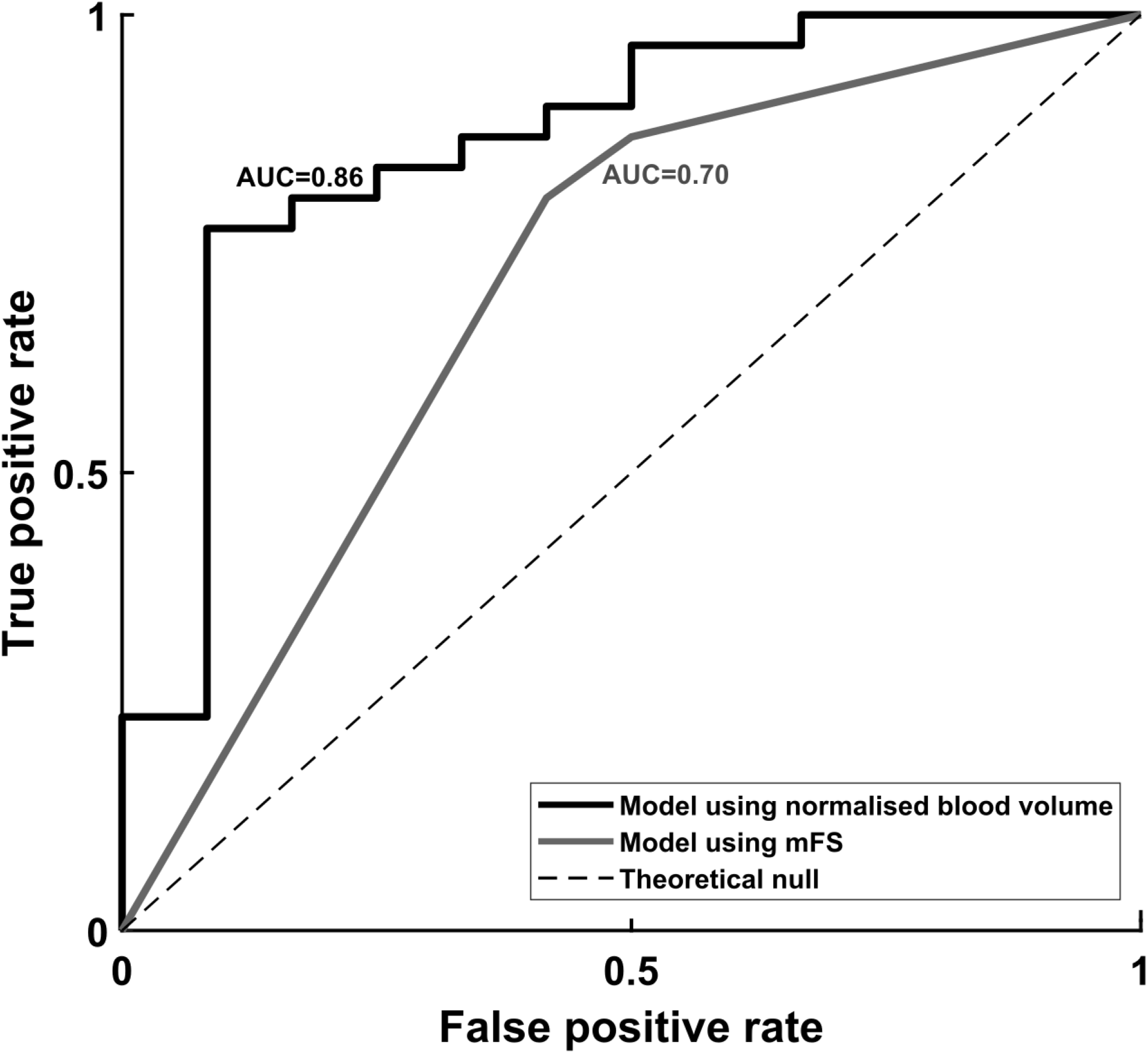
Receiver operating characteristic (ROC) curve demonstrating the performance characteristics of the binary classifier fit in the logistic regression model. Black = univariate logistic regression model using normalised total blood volume; grey = logistic regression model using dummy-coded modified Fisher score values.

**Table 2.** Logistic regression fit parameters for all variables in the full predictive model including confounding variables. Following inclusion of potential confound variables into the logistic regression against radiological vasospasm, the only significant predictor remained the estimate for blood volume (*, p < .05, italic typeface). Note that estimates for logistic regression are given in the form of log odds. χ^2^-statistic vs. constant model: F = 20.6, p-value = 0.00215.

|  | Estimate (log odds) | Confidence interval | t value | p value |
| --- | --- | --- | --- | --- |
| **Intercept** | 2.3975 | [-4.40, 9.20] | 0.69 | 0.49 |
| **Age** | -0.099725 | [-0.22, 0.02] | -1.69 | 0.09 |
| **Gender** | 1.4887 | [-0.57, 3.54] | 1.42 | 0.16 |
| **Treatment: coiled** | 0.14742 | [-2.03, 2.33] | 0.13 | 0.89 |
| **EVD inserted** | 0.78481 | [-1.43, 3.00] | 0.70 | 0.49 |
| **Lumbar drain inserted** | 0.08629 | [-2.26, 2.43] | 0.072 | 0.94 |
| ***Normalised blood volume\**** | *0.0741* | *[0.0077, 0.1406]* | 2.19 | *0.03* |

For comparison, a logistic regression model using only mFS scores was also fitted (see Supplementary Results Table S1). For dummy coding, mFS scores 1 and 2 were combined. While this also outperformed a constant model (χ^2^ = 6.31; p < .043), it had less predictive ability (AUC = 0.70) as compared to the previous models described (Figure 4).

The mFS was significantly associated with larger volumes of subarachnoid blood (normalised blood volume for mFS 1: 8.5±5.7 cm^3^; mFS 2: 23.6±12.8 cm^3^; mFS 3: 53.9±37.8 cm^3^; mFS 4: 61.3±29.0 cm^3^; one-way ANOVA: F = 7.35, p < .001). Following Bonferroni correction for multiple comparisons, scans of mFS grade 4 contained significantly larger blood volume than scans of mFS grade 1 (p = .001) and grade 2 (p = .037). To assess whether blood volume contained additional independent information, it was added to a logistic regression model containing mFS alongside confounding variables (see Tables 2, S2, S3 for full models). In doing so, model predictive power was improved (Likelihood ratio = 8.00; p < .0047) and predictive error was reduced (mFS only: AIC = 47.5, BIC = 58.0; mFS and blood volume: AIC = 41.55, BIC = 53.7). However, when the mFS was added to the blood volume model, no increase in predictive power was shown (Likelihood ratio = 1.94; p = 0.16). Similarly, leave-one-out cross validation of the full model demonstrated a classification accuracy of 78.6%, greater than the classification error for models containing blood volume (71.4%) or mFS (69.0%) alone. Taken together, these results indicate that the normalised blood volume contains additional information to the mFS, and constitutes a significant and independent predictor of radiological vasospasm.

### Greater subarachnoid blood volume is associated with worse vasospasm severity

To investigate whether segmented blood load was associated with the severity of radiological vasospasm, we extracted the subjective impression of severity from radiologist reports. A one-way ANOVA demonstrated a significant association between normalised blood volume and subjective severity (F = 5.42; p = 0.003). Post-hoc testing using the Tukey-Kramer method for multiple comparisons demonstrated that patients with reported ‘moderate’ and ‘severe’ radiological vasospasm had significantly greater blood load than those without vasospasm (p = 0.028 and p = 0.004, respectively; Figure 5A). However, no significant difference in blood volume was found between patients with mild, moderate, and severe vasospasm (p > .05 throughout).

**Fig 5:**
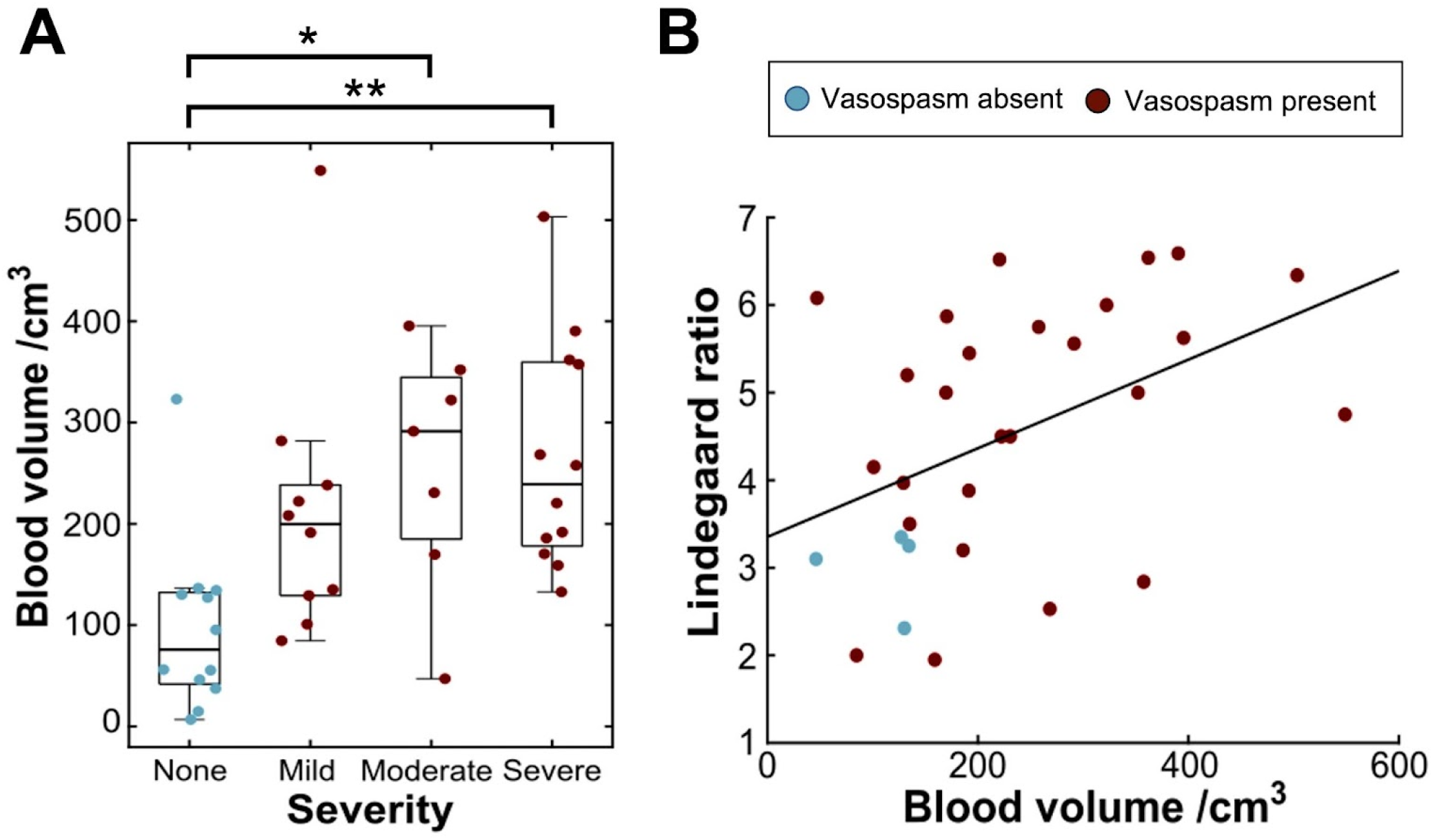
Associations between subarachnoid blood volume and metrics of severity of radiological vasospasm. A: Boxplots of blood volume grouped by radiologist’s impression of subjective severity of vasospasm. B: Scatter plot showing the greatest recorded Lindegaard ratio from TCD plotted against normalised blood volume. (* = p <.05, ** = p<.01)

To further probe this result, we evaluated quantitative metrics of vasospasm severity (Figure 5B). We extracted the largest Lindegaard ratio observed for all patients who received documented TCDs (n = 30 patients received at least one TCD positive or negative for vasospasm following admission). Similarly, blood volume was significantly correlated with higher Lindegaard ratios (r = 0.45; p = 0.014), indicating that blood load was related to the severity of radiological changes seen in vasospasm-positive patients. This relationship remained significant when only TCD-positive vasospasm patients were included (n=23; r = 0.46; p = 0.027).

### Subarachnoid blood volume influences the duration and frequency of vasospasm episodes

Of patients who developed vasospasm, radiological evidence of vasospasm was first reported an average of 5.30±3.41 days after presentation, and persisted for 3.62±3.22 days, consisting of 1.29±1.22 discrete episodes.

Increased blood volume was significantly associated with more discrete episodes of radiological vasospasm (r = 0.57, p < .001) [Figure 6A]. Similarly, there was a strong association between blood volume and vasospasm duration (r = 0.54, p < .001; Figure 6B). However, no association was found between subarachnoid blood volume and the time from admission date to first vasospasm (r = -0.13; p = 0.51).

**Fig 6:**
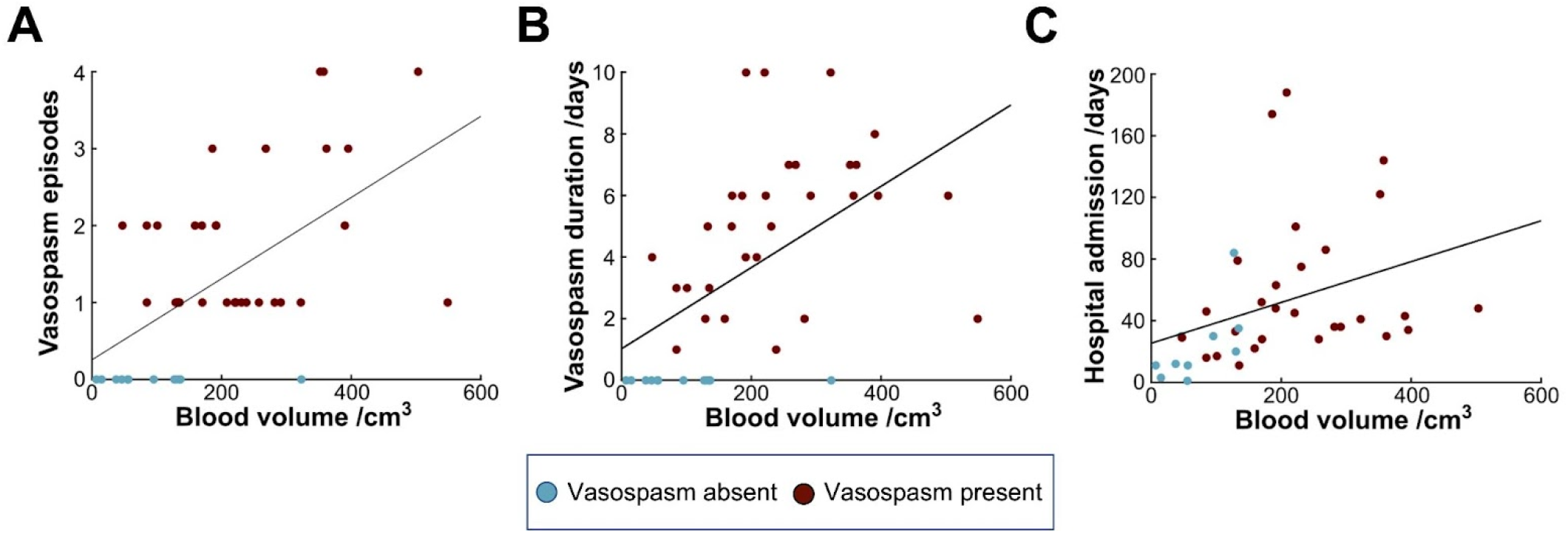
Associations between subarachnoid blood volume and temporal vasospasm-related outcomes. A: Scatter plot showing the number of discrete episodes of vasospasm against normalised blood volume. B: Scatter plot showing the duration of vasospasm in days plotted against normalised blood volume. C: Scatter plot showing the total length of hospital admission plotted against normalised blood volume.

### Association of subarachnoid blood volume with neurosurgical patient outcomes

Blood volume was not significantly associated with length of ITU stay (r = 0.16; p = 0.34), and there was no difference in mean blood volume between patients who subsequently died and those who did not, although this approached significance (fatality group: 77.9±43.9 cm^3^; non-fatality group: 47.0±30.2 cm^3^; t = 1.87; p = 0.07). However, blood volume was significantly correlated with total length of hospital admission (r = 0.36; p = 0.027; Figure 6C), indicating that patients with larger subarachnoid blood load may experience a more complicated or prolonged recovery.

## Discussion

### Summary

In this article, we present a novel ITK-SNAP-based pipeline for reliable and efficient segmentation of subarachnoid blood on initial CT head scans using semi-automated methods and expertly verified. Utilising this framework, we show that total segmented blood volume following subarachnoid haemorrhage is associated with several vasospasm-relevant outcomes. We show that greater total blood volume is significantly associated with a greater probability of subsequent radiological vasospasm, with the odds of vasospasm occurring increasing by approximately 7% for each cm^3^ of subarachnoid haemorrhage. We also demonstrate that subarachnoid blood load influences the natural history of vasospasm both in terms of duration and number of distinct episodes, and is associated with overall hospital length of stay.

### Interpretation and context

Our pipeline successfully produced segmentations of blood from CT head scans for all scans included in the study. Segmentations were close to ground truth as defined by corrected segmentations provided by a consultant neurovascular surgeon, indicating that our methods were accurately and reliably delineating blood from other tissue, in spite of its complex and tortuous morphology and also in spite of a wide array of scanner acquisition protocols from several referring hospitals. While previous machine learning methods have been used to detect and classify intracranial haemorrhages[36,37], by providing saliency maps highlighting probable regions where blood is distributed[38], these methods do not produce segmentations from which precise blood volumes can be obtained. Quantitative volumetric segmentations have typically been applied to haemorrhagic lesions from traumatic brain injuries, with focus on subdural haematoma, extradural haematoma, and intraparenchymal haemorrhage[39–43]. Less work has attempted the automated segmentation of subarachnoid blood[37], and to our knowledge this work represents the first use of machine learning techniques to segment blood from CT head scans in aneurysmal SAH patients.

Our results demonstrate that the initial blood burden following subarachnoid haemorrhage has future consequences. Previous literature has focused on predicting the risk of symptomatic vasospasm (or delayed cerebral ischaemia) based on blood volume on CT head scans[4,7,8,11,12], with few papers addressing angiographic or radiological features. In our cohort, haemorrhagic blood load was associated with greater radiological vasospasm risk, episodes, duration, severity and a longer length of stay in hospital. Radiological vasospasm is itself well known to be strongly associated with delayed cerebral ischaemia and poorer functional outcomes[21,22].

That we found both blood volume and the radiological vasospasm to be significantly associated with overall hospital admission length, suggests that subarachnoid blood burden and its sequelae may complicate and prolong hospital admissions. This may be because of additional complications associated with higher blood volumes including hydrocephalus that in turn require invasive treatment. Although the length of intensive care stay was not associated with blood volume in our cohort, we did note an association with greater mortality that approached significance.

To address the clinical utility of our segmentations, we compared the predictive power of total subarachnoid blood volume with that of the modified Fisher scale, a qualitative grading metric commonly used in clinical practice. The modified Fisher scale alone possesses a number of limitations. Recent work has highlighted its inherent subjectivity, demonstrating only moderate inter-rater reliability scores[15]. Further, its predictive power is limited by its qualitative nature, resulting in a low-dimensional and low-resolution description of blood load and distribution. In our dataset, the modified Fisher scores also significantly predicted vasospasm risk but with reduced accuracy. In addition, we found that normalised blood volume provides additional information to the logistic regression model that is independent of the modified Fisher scale, and therefore may be incorporated into the future development of radiological vasospasm risk scales. Nonetheless, it may be that information about blood volume distribution across compartments (i.e. cisternal and ventricular blood) is provided the modified Fisher scale, and this would not be captured by a total blood volume value. However, as blood segmentations can be further extended to include information about the spatial distribution of blood in the brain, we suggest that total blood volume provides a potential powerful regressor for predicting vasospasm.

### Limitations and strengths

Our interpretations are limited by the modest number of patients included, alongside the retrospective design of the study. Furthermore, the statistics presented here are exploratory, although the significance of the associations presented remained so after multiple comparison corrections. Additionally, we describe several steps to demonstrate that regression models fitted were robust. Although our fitted and internally validated regression model demonstrates good performance on our single-centre dataset, further work with larger, multi-centre datasets will be required to cross-validate and confirm the findings reported above. However, the results nonetheless highlight important and unexplored potential areas of further study within the vasospasm literature, and our presented analysis pipeline can easily be extended to larger datasets, prospective studies, collaborative segmentations, and more sophisticated statistical models.

The modified Fisher score was validated for prediction of delayed cerebral ischaemia[8] rather than radiological vasospasm, and so may not be expected to predict radiological vasospasm more accurately than blood volume. DCI remains challenging to diagnose, especially in sedated or high-grade patients[44], and unsurprisingly its onset is often missed[23]. However, early angiographic vasospasm is significantly associated with the subsequent development of DCI[45,46]. Further, vasospasm as detected on TCD or CTA is well correlated with clinical deficits[21,22,45,46], and both vasospasm duration and severity can be used to assess the likelihood of delayed cerebral ischaemia and therefore the need for clinical interventions such as angioplasty[21,24,47,48]. Therefore, we propose that radiological detection of vasospasm, as an objective biomarker, may provide important information that may guide clinical decision making. Total blood volume provides useful information that can be used to predict the likelihood of significant vasospasm, and identify a subpopulation of patients that may require more stringent monitoring and/or intervention. Further modelling would be necessary to determine the influence of CSF drainage factors which may assist in reducing subarachnoid blood volume.

Fully manual segmentations drawn out by experts or trained raters, despite being considered the gold standard method, are time- and labour-intensive, often require lengthy training periods[49], and risk introducing substantial intra-rater and inter-rater variability and bias arising from various sources, including differences in opinion about the ground truth segmentation and in anatomical knowledge about relevant structures[50]. Accordingly, previous work has shown substantial variation in consistency and reproducibility of manual segmentations across both raters and structures[51–53]. Over recent years, semi-automated and automated methods have begun to match and even outperform manual segmentation in metrics of precision and inter-rater variability across a variety of modalities and structures[49,52,54,55].

Our semi-automated method adds to the growing literature of potential applications for machine learning methods in radiological interpretation and triage, and removes some of this intra- and inter-rater variability. Many computer-assisted methods for delineation of blood volume have previously focused on segmentation of haemorrhages within non-subarachnoid space [39,56–59]. However, application of these methods to aSAH has been noted to be challenging [40,57], and accordingly Dice scores for segmentations of subarachnoid blood have been consistently lower than for other haemorrhage subtypes [37,53,60,61], and convolutional networks used to automatically segment intracranial haemorrhage that includes subarachnoid blood have only achieved low-to-moderate Dice scores [62,63].

We attempted to mitigate any bias on behalf of the rater through expert assessment and correction, and correspondingly our mean Dice score between original and corrected segmentation was high (0.92), indicating excellent agreement between rater segmentation and expert opinion that required minimal correction. This score was substantially larger than those comparing manual segmentations by different observers [53,61], and in previous literature [37,53,60,61]. In particular, Boers et al. [53] previously utilised similar methods to segment aSAH, but achieved only moderate Dice scores for its automated segmentations (mean Dice score 0.55, range 0.00 - 0.83). Nonetheless, some variability remains, as labelling training data for the classifier and placement of seeds remain as manual steps that may lead to unintentional biases in volumes.

Segmentations took on average 10-15 minutes per scan for the rater to perform. Although this is notably faster than manual segmentations, this remains a non-trivial period of time that may bottleneck scan reporting, and may therefore be unfeasible in clinical radiology contextS.

Potential extensions to a fully automated pipeline (such as nnU-Net[64], which has already seen use in brain tumour segmentation [65,66]) would address a number of these limitations and allow for development of a clinically valuable toolkit. Validated, fully automated segmentations would allow for faster, reproducible and accurate delineation of blood distribution and volume, and may remove variability introduced in manual steps. Nonetheless, these methods still require training and validation, and this dataset serves as an important repository in facilitating this research.

## Conclusion

The semi-automated pipeline presented here robustly segments fresh blood from CT head scans on admission following subarachnoid haemorrhage. Our segmentation pipeline is significantly faster than manual segmentations, and demonstrates high accuracy when compared with expertly corrected volumes. Using these methods, we demonstrate that blood load following aSAH is associated with risk and timeline of radiological vasospasm. Notably, the odds of developing radiological vasospasm were greater for larger haemorrhage volumes, with a 7% increase in vasospasm odds per additional cm^3^ of blood on the scan, and observed vasospasm is likely to be more severe and persist for longer. Total blood volume constitutes an independent predictor for radiological vasospasm from the clinically employed modified Fisher scale, and carries potential for extension in the future to fully automated segmentation pipelines, and for the development of more sophisticated radiological risk scores for vasospasm.

## Supporting information

Supplementary Methods and Results

## Data Availability

Due to the sensitive nature of the data included in this research, supporting imaging data are not available. Analysis code and pipelines are available from the corresponding author upon reasonable request.

## Abbreviations List

AIC: Akaike information criterion
AUC: area-under-curve
aSAH: aneurysmal subarachnoid haemorrhage
BET: brain extraction tool
BIC: Bayesian information criterion
CT: computed tomography
CTA: computed tomography angiography
CTH: computed tomography (of head)
DICOM: digital imaging and communications in medicine
DCI: delayed clinical ischaemia
DSA: digital subtraction angiography
EHR: electronic healthcare record
EVD: external ventricular drain
HU: Hounsfield unit
ITU: intensive therapy unit
IVH: intraventricular haemorrhage
mFS: modified Fisher scale
PACS: picture archiving and communication system
ROC: receiver operating characteristic
TCD: transcranial Doppler

