## Supplementary Methods and Results for "Predicting vasospasm risk using first presentation aneurysmal subarachnoid haemorrhage volume: a semi-automated CT image segmentation analysis in ITK-SNAP"

### Supplementary Material

#### Supplementary Methods

##### Data and statistical analysis

The Hosmer-Lemeshow test looks for significant differences in observed versus predicted outcomes of a model across its deciles of fitted risk values, whereas the Stukel test adds additional predictors derived from the fitted values of the model and tests for significance of these predictors, indicating poor fit. Both detect departures from linearity and unspecified interactions, and are well discussed elsewhere [1,2].

#### Supplementary Figures and Tables

**S1 Table: Logistic regression fit parameters using the modified Fisher score only.** Modified Fisher scores were dummy-coded before inclusion in the model, with mFS 1-2 grouped together as the referent. Note that estimates for logistic regression are given in the form of log odds.

| | | **Estimate (log odds)** | **Confidence interval** | ***t* value** | ***p* value** |
|---|---|---|---|---|---|
| **Intercept** | | -0.40547 | [-1.6706, 0.8597] | 0.62814 | 0.53 |
| **Modified Fisher score** | **1-2** | Referent | | | |
| | **3** | 1.0986 | [-3.1190, 2.3080] | 0.79354 | 0.43 |
| | ***4**** | *1.9741* | *[0.3838, 3.5644]* | *2.433* | *0.015* |

42 observations, 39 error degrees of freedom. Dispersion: 1
Chi^2-statistic vs. constant model: 6.31, p-value = 0.0426

**S2 Table: Logistic regression fit parameters using the mFS scores and confounding variables.** Modified Fisher scores were dummy-coded before inclusion in the model, with mFS 1-2 grouped together as the referent. Note that estimates for logistic regression are given in the form of log odds.

| | | **Estimate (log odds)** | **Confidence interval** | ***t* value** | ***p* value** |
|---|---|---|---|---|---|
| **Intercept** | | 2.8839 | [-2.3849, 8.1527] | 1.0728 | 0.28 |
| **Age** | | -0.08264 | [-0.1796, 0.0143] | -1.6702 | 0.09 |
| **Gender** | | 1.6176 | [-0.4870, 3.7221] | 1.5065 | 0.13 |
| **Treatment: coiled** | | -0.63353 | [-1.8616, 0.5945] | -1.0111 | 0.31 |
| ***EVD inserted**** | | *2.2294* | *[0.0721, 4.3867]* | *2.0255* | *0.04* |
| **Lumbar drain inserted** | | 0.12393 | [-2.011, 2.2490] | 0.11431 | 0.91 |
| **Modified Fisher score** | **1-2** | Referent | | | |
| | **3** | -0.53881 | [-3.8684, 2.7908] | -0.31718 | 0.75 |
| | **4** | 1.4268 | [-0.6977, 3.5513] | 1.3164 | 0.19 |

42 observations, 34 error degrees of freedom. Dispersion: 1
Chi^2-statistic vs. constant model: 14.7, p-value = 0.0399
AIC: 41.5505, BIC: 57.9723, AUC: 0.8583

**S3 Table: Logistic regression fit parameters using the blood volume, mFS scores, and confounding variables.** Modified Fisher scores were dummy-coded before inclusion in the model, with mFS 1-2 grouped together as the referent. Note that estimates for logistic regression are given in the form of log odds.

| | | Estimate (log odds) | Confidence interval | *t* value | *p* value |
|---|---|---|---|---|---|
| **Intercept** | | 3.7773 | [-3.3313, 10.8859] | 1.0415 | 0.30 |
| **Age** | | -0.11272 | [-0.2398, 0.0144] | -1.7378 | 0.08 |
| **Gender** | | 1.7815 | [-0.4559, 4.0188] | 1.5606 | 0.12 |
| **Treatment: coiled** | | -0.75851 | [-2.3043, 0.7873] | -0.96175 | 0.34 |
| **EVD inserted** | | 0.9748 | [-1.3261, 3.2757] | 0.83036 | 0.41 |
| **Lumbar drain inserted** | | 0.020753 | [-2.6246, 2.6661] | 0.015376 | 0.99 |
| ***Normalised blood volume**** | | *0.076197* | *[0.0008, 0.1516]* | *1.9798* | *0.048* |
| **Modified Fisher score** | **1-2** | Referent | | | |
| | **3** | -3.0509 | [-7.7477, 1.6459] | -1.2732 | 0.20 |
| | **4** | –0.27451 | [-2.9733, 2.4243] | -0.19936 | 0.84 |

42 observations, 33 error degrees of freedom. Dispersion: 1
Chi^2-statistic vs. constant model: 22.7, p-value = 0.00377
AIC:  41.5505, BIC: 53.7142, AUC: 0.9111
